# Seroprevalence of anti-SARS-CoV-2 IgG antibodies in Juba, South Sudan: a population-based study

**DOI:** 10.1101/2021.03.08.21253009

**Authors:** Kirsten E. Wiens, Pinyi Nyimol Mawien, John Rumunu, Damien Slater, Forrest K. Jones, Serina Moheed, Andrea Caflish, Bior K. Bior, Iboyi Amanya Jacob, Richard Lino Loro Lako, Argata Guracha Guyo, Olushayo Oluseun Olu, Sylvester Maleghemi, Andrew Baguma, Juma John Hassen, Sheila K. Baya, Lul Deng, Justin Lessler, Maya N. Demby, Vanessa Sanchez, Rachel Mills, Clare Fraser, Richelle C. Charles, Jason B. Harris, Andrew S. Azman, Joseph F. Wamala

## Abstract

**Background:** Relatively few COVID-19 cases and deaths have been reported through much of sub-Saharan Africa, including South Sudan, although the extent of SARS-CoV-2 spread remains unclear due to weak surveillance systems and few population-representative serosurveys.

**Methods:** We conducted a representative household-based cross-sectional serosurvey in Juba, South Sudan. We quantified IgG antibody responses to SARS-CoV-2 spike protein receptor-binding domain and estimated seroprevalence using a Bayesian regression model accounting for test performance.

**Results:** We recruited 2,214 participants from August 10 to September 11, 2020 and 22.3% had anti-SARS-CoV-2 IgG titers above levels in pre-pandemic samples. After accounting for waning antibody levels, age, and sex, we estimated that 38.5% (32.1 - 46.8) of the population had been infected with SARS-CoV-2. For each RT-PCR confirmed COVID-19 case, 104 (87-126) infections were unreported. Background antibody reactivity was higher in pre-pandemic samples from Juba compared to Boston, where the serological test was validated. The estimated proportion of the population infected ranged from 30.1% to 60.6% depending on assumptions about test performance and prevalence of clinically severe infections.

**Conclusions:** SARS-CoV-2 has spread extensively within Juba. Validation of serological tests in sub-Saharan African populations is critical to improve our ability to use serosurveillance to understand and mitigate transmission.

## Background

Globally, over 100 million cases and over 2 million deaths have been attributed to Coronavirus disease 2019 (COVID-19) as of February 16, 2021 [1]. The majority of the cases have been reported from Europe and the Americas where the pandemic has overwhelmed some of the best health systems. In Africa, over 2.7 million cases and nearly 70,000 deaths have been reported [1]. The reasons for the generally lower prevalence and mortality associated with COVID-19 in Africa, particularly during the first 6-8 months of the pandemic, are unclear, but may be due to differences in age distribution, immune history, climate, mitigation measures, or conditions for spread, such as travels and connectivity between geographic regions [2,3]. However, the true spread of Severe acute respiratory virus coronavirus 2 (SARS-CoV-2) has been underestimated due to limited testing capabilities, under-reported deaths, and undetected mild and asymptomatic infections. Population-based serological surveys that measure anti-SARS-CoV-2 antibodies can help shed light on this by estimating the extent of infections [4]. Hundreds of serosurveys have been conducted worldwide to estimate SARS-CoV-2 seroprevalence [5] and as of February 23, 2021, only 15 of the studies published or available in pre-print were conducted in sub-Saharan Africa [6–20], of which 2 were population-based and representative studies (in Nigeria and Ethiopia). No serosurveys have been conducted in South Sudan.

South Sudan confirmed its first case of COVID-19 in the capital of Juba on April 4, 2020 [21], and saw its first wave of reported cases from May through July of 2020 (Figure 1). By August 31, 2020, a total of 1,873 virologically confirmed SARS-CoV-2 infections (∼47 per 10,000 population) had been reported in Juba out of 18,156 RT-PCR tests conducted. RT-PCR testing in South Sudan, including Juba, has remained limited due to scarce reagents, few testing sites, limited willingness to be tested, and logistic challenges. Thus, like much of sub-Saharan Africa, the true extent of SARS-CoV-2 spread in the population remains unknown. Understanding spread can inform cost-benefit analyses of mobility restrictions, lockdown measures, and other public health interventions. In sub-Saharan Africa, it is important to weigh the decrease in transmission against the socio-economic impact of these measures, since this can translate into excess mortality due to food insecurity and constrained access to basic services.

**Figure 1.**
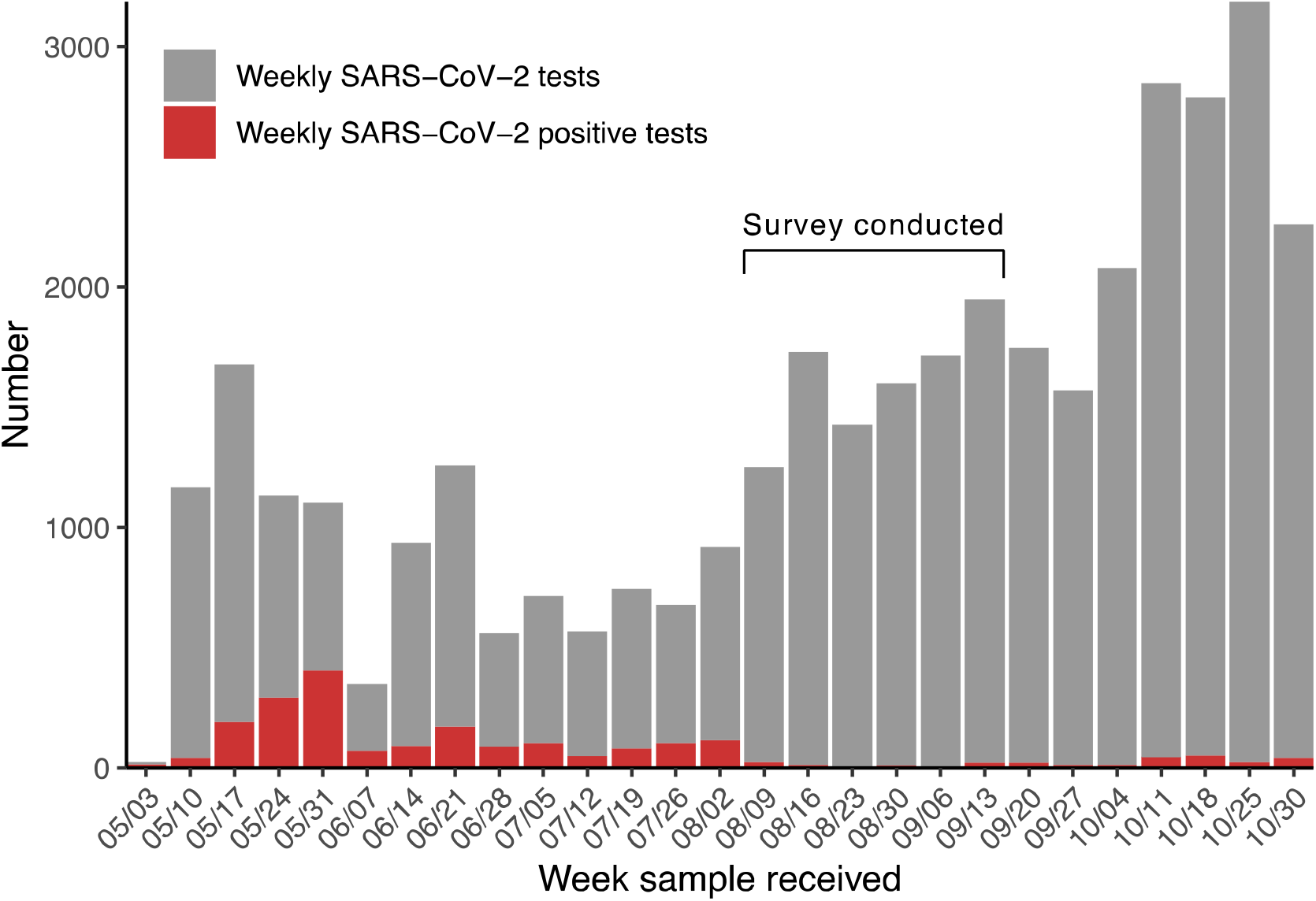
Number of weekly SARS-CoV-2 tests and COVID-19 cases reported in Juba. Weekly SARS-CoV-2 PCR tests performed from the week of May 3, 2020 to the week of October 30, 2020 in Juba. Grey bars show number of tests conducted per week, and red bars show the number of those tests that were positive for SARS-CoV-2. The first COVID-19 case was identified on April 2 and confirmed on April 4, 2020 [21].

Understanding the extent of SARS-CoV-2 spread is particularly important for guiding COVID-19 mitigation efforts in light of South Sudan’s complex humanitarian and public health context. South Sudan has experienced years of conflict and continues to undergo a grade three protracted crisis as defined by the World Health Organization [22]. The crisis has led to 1.61 million internally displaced persons (IDP), severe food insecurity affecting half the population (6 million), and 1.3 million malnourished children [22,23]. In Juba, 28.7% of households indicated that they were unable to access health care services when needed in the first six months of the pandemic, and this increased to 43.2% in the lowest wealth quintile [24]. The majority mentioned that the cost of healthcare was the main barrier [24]. These underlying vulnerabilities may increase risk of SARS-CoV-2 spread and may themselves be compounded by the direct and indirect effects of the epidemic.

In order to estimate the seroprevalence of anti-SARS-CoV-2 antibodies and associated risk factors in Juba, we conducted a representative household-based cross-sectional serosurvey. Here, we present the results of this serosurvey and discuss their implications for SARS-CoV-2 surveillance in South Sudan, as well as more broadly for serological studies conducted in Africa and worldwide.

## Methods

### Study design and participants

We conducted a cross-sectional serosurvey in residential neighborhoods of the City of Juba and Juba County following protocols from the World Health Organization’s Unity Studies [4]. The urban extent was mapped based on built-up areas, local administrative boundaries, and the existing transport network within the payams (i.e. fourth administrative divisions) of Northern Bari, Munuki, Juba, Kator, Rejaf, and Gondokoro. Juba IDP Camp I and III, the former UNMISS Protection of Civilians (PoC) sites, were not included in the sampling frame.

The survey employed two-stage cluster sampling. Enumeration areas (EAs) were used as clusters and were based on building footprints derived from high-resolution satellite imagery and field mapping of non-residential areas. The target sample size was 2,750 (50 clusters of 55 respondents each). EAs were selected using probability proportional to size sampling. Three sampled EAs inhabited by families of military personnel were randomly replaced due to denied access. Within each sampled EA, 11 residential structures were randomly selected as households to recruit into the study.

Households were defined as a group of individuals that sleep under the same roof most nights and share a cooking pot. All household members were eligible for inclusion if they or their guardian provided written consent to participate, were at least 1 year of age, and had lived in the area at least 1 week before the survey, regardless of current or past illness. For households with more than 10 people, only the first-degree relatives of the head of household were eligible for study inclusion.

Eligible participants were interviewed to collect information about sociodemographic characteristics, history of respiratory illness, history of SARS-CoV-2 tests, potential exposure risks in the previous two weeks, household deaths, and COVID-19 prevention measures. Dried blood samples (DBS) were collected by applying a few drops of blood, drawn by lancet from the finger, heel, or toe, onto Whatman 903 (Whatman plc, Springfield Mill, UK), Ahlstrom grade 226 (Ahlstrom Corporation, Helsinki, Finland) filter paper. Blood was allowed to thoroughly saturate the paper and air dried overnight at an ambient temperature (median temperature□=□31°C, median humidity□=□33%). DBS were stored in low gas-permeability plastic bags with desiccant added to reduce humidity. DBS were transported at ambient temperature to Massachusetts General Hospital in Boston, USA following IATA protocol and stored at 4°C until tested. The study protocol was approved by South Sudan Ministry of Health Ethics Review Board.

### Laboratory analysis

DBS were eluted and tested for the presence of anti-SARS-CoV-2 immunoglobulin G (IgG) antibodies targeting the receptor-binding domain (RBD) of the spike protein of SARS-CoV-2 using a quantitative enzyme-linked immunosorbent assay (ELISA) previously developed and validated at Massachusetts General Hospital [25]. This assay quantifies RBD-specific antibody concentrations (μg/mL) using IgG-specific anti-RBD monoclonal antibodies, and the full protocol used for eluting DBS samples for the ELISA are detailed online [26]. Validation of this test was originally based on PCR-positive infections and pre-pandemic samples from Boston, USA. To help decide on an appropriate positivity threshold and assess assay specificity, we measured background antibody reactivity using 104 dried blood spot samples collected in Juba in 2015 [27]. We then selected a seropositivity threshold (0.32 μg/mL) that corresponded to 100% specificity in these pre-pandemic samples (i.e. their highest value, Supplementary Figure 1) and 99.7% in the pre-pandemic samples collected from the USA.

### Statistical analysis

To estimate test sensitivity, we used data from a cohort of mild and severe confirmed SARS-COV-2 infections in Boston whose antibody levels had been characterized at multiple time points post symptom onset [25], and supplemented these with recent data collected by dried blood spot from non-hospitalized PCR-positive individuals in Boston (Supplementary Figure 2). Based on the trends in positive RT-PCR results in Juba, we assumed that most serosurvey participants, if previously infected, would have been exposed to SARS-CoV-2 more than 30 days before the survey (Figure 1), and restricted the positive-control data to observations more than 30 days post-symptom onset. As infections with mild disease may lead to lower levels of detectable antibodies [28], we created a synthetic cohort of positive controls such that 80% of the sample was from mild infections (defined as not needing hospitalization in the Boston health system), and 20% severe cases (defined as hospitalized, excluding deaths), consistent with previous analyses [29,30] and the predominantly young population in Juba [31]. To evaluate the impact of this assumption, we performed sensitivity analyses testing a range of assumed mild case fractions (60 - 100%) in the positive control dataset.

To estimate the proportion of the population previously infected (referred to as seroprevalence), we followed a previously published Bayesian approach [32] using a regression model that accounted for age and sex of the study population integrated with a binomial model of the sensitivity and specificity of the ELISA. This approach allowed us to adjust the estimates for test performance, while propagating uncertainty around test performance in the adjusted estimates. We implemented the models in the Stan probabilistic programming language [33] using the *rstan* package in R. We post-stratified our modeled results accounting for the age distribution of urban populations in South Sudan [31] in order to generate population-representative seroprevalence estimates. Unless otherwise indicated, we reported estimates as the mean of the posterior samples and 95% Credible Interval (CrI) as the 2.5^th^ and 97.5^th^ percentiles of this distribution.

In addition, we calculated the relative risk of being seropositive within age and sex subsets using the posterior draws for each regression coefficient. We also estimated relative risk of being seropositive among non-working adults compared to working adults, children, and students using a log-binomial regression model. We estimated implied infections by multiplying estimated seroprevalence by 510,000, Juba’s estimated population size [34]. We then estimated the ratio of reported to unreported infections by subtracting PCR confirmed COVID-19 cases in Juba as of August 31, 2020 from total implied infections and dividing this estimate of unreported infections by RT-PCR confirmed COVID-19 cases. Analysis code is available online (https://github.com/HopkinsIDD/juba-sars-cov-2-serosurvey), and additional methodological details can be found in the Supplementary methods.

## Results

A total of 2,214 participants between 1 and 84 years of age from 435 households were recruited and provided dried blood spot samples between August 10 and September 11, 2020. Of these, 1,840 (83.2%) had complete interview and demographic data available and 374 were missing interview data due to data collection device failures and data entry issues. Based on these 1,840 participants, over half (62.4%) were female and 73.5% were between 10 and 49 years of age (Table 1), consistent with the predominantly young population in South Sudan [31].

**Table 1.** Characteristics of participants with interview data available (n = 1840). N is total number of participants included in each category and % indicates percentage of the participants that fell within each category.

| Characteristic | Group | N | % |
| --- | --- | --- | --- |
| Sex | Female | 1149 | 62.4 |
|  | Male | 691 | 37.6 |
| Age | 1 - 4 years | 68 | 3.7 |
|  | 5 - 9 years | 224 | 12.2 |
|  | 10 - 19 years | 448 | 24.3 |
|  | 20 - 49 years | 905 | 49.2 |
|  | 50 - 64 years | 120 | 6.5 |
|  | > 65 years | 75 | 4.1 |
| Payam | Northern Bari | 788 | 42.8 |
|  | Juba | 141 | 7.7 |
|  | Muniki | 397 | 21.6 |
|  | Kator | 229 | 12.4 |
|  | Rejaf | 135 | 7.3 |
|  | Gondokoro | 150 | 8.2 |
| Occupation | None | 408 | 22.2 |
|  | Child | 386 | 21.0 |
|  | Student | 388 | 21.1 |
|  | Market merchant | 89 | 4.8 |
|  | Healthcare worker | 12 | 0.7 |
|  | Taxi driver | 16 | 0.9 |
|  | Farmer | 164 | 8.9 |
|  | Working with animals | 10 | 0.5 |
|  | Civil servant | 120 | 6.5 |
|  | Health laboratory worker | 2 | 0.1 |
|  | Teacher | 20 | 1.1 |
|  | Traditional healer | 1 | 0.1 |
|  | Religious leader | 8 | 0.4 |
|  | Other | 216 | 11.7 |
| Reported test for SARS-CoV-2 | No | 1816 | 98.7 |
|  | Yes | 22 | 1.2 |
|  | Unknown | 2 | 0.1 |
| Reported SARS-CoV-2 test result | Negative | 15 | 0.8 |
|  | Positive | 5 | 0.3 |
|  | Unknown | 2 | 0.1 |

We found that 22.3% (494/2214) of samples collected during the survey were above the test positivity threshold, which we selected to have 100% specificity against pre-pandemic samples from Juba. However, this crude seroprevalence estimate does not account for test sensitivity.

We estimated that test sensitivity was 64.7% based on a cohort of PCR-confirmed COVID-19 cases in the USA [25] and a series of assumptions about time since infection and the prevalence of mild infections in Juba (see Methods). Using the samples with interview and matched demographic data available, we estimated an adjusted seroprevalence of 38.5% (95% Credible Interval [CrI], 32.1 - 46.8) in August 2020. Seroprevalence in the matched dataset was nearly indistinguishable from the full dataset (Supplementary Table 1), thus we used the matched dataset in all subsequent analyses. These results imply that for each RT-PCR confirmed COVID-19 case tested by the end of August, 104 (95% CrI 87 - 126) SARS-CoV-2 infections were unreported. We found no difference in the risk of seropositivity by sex (Table 2). Though, we found that risk of seropositivity was lowest among participants 20 to 49 years old (Table 2) and that adjusted seroprevalence in this group was 32.1% (95% CrI 25.5 - 39.6) (Table 2). Seroprevalence was highest among individuals 10 to 19 years old at 45% (95% CrI 36.3 - 55.5) (Table 2). Non-working adults had 35% lower risk (relative risk=0.65, 95% Confidence Interval 0.50 - 0.82) of being seropositive compared to working adults, children, and students.

**Table 2.** Crude seropositivity, adjusted seroprevalence, and relative risk of seropositivity by age and sex following Stringhini and colleagues [32]. Data are n (%) unless otherwise indicated. Ages 20 to 49 years and female are the reference groups.

| Category | N | Test positive | Test negative | Seroprevalence<br>(95% CrI) | Relative risk<br>(95% CrI) |
| --- | --- | --- | --- | --- | --- |
| 1 - 4 years | 68 | 20 (29.4%) | 48 (70.6%) | 44.6 (32.5 - 57.8) | 1.40 (1.04-1.79) |
| 5 - 9 years | 224 | 52 (23.2%) | 172 (76.8%) | 39.3 (29.2 - 50.9) | 1.23 (0.96-1.53) |
| 10 - 19 years | 448 | 124 (27.7%) | 324 (72.3%) | 45 (36.3 - 55.5) | 1.41 (1.19-1.67) |
| 20 - 49 years | 905 | 167 (18.5%) | 738 (81.5%) | 32.1 (25.5 - 39.6) | -- |
| 50 - 64 years | 120 | 31 (25.8%) | 89 (74.2%) | 43 (30.4 - 58.4) | 1.34 (0.98-1.75) |
| 65 - 84 years | 75 | 17 (22.7%) | 58 (77.3%) | 38.8 (25.7 - 55) | 1.22 (0.82-1.68) |
| Female | 1149 | 260 (22.6%) | 889 (77.4%) | 32.1 (25.5 - 39.6) | -- |
| Male | 691 | 151 (21.9%) | 540 (78.1%) | 30.4 (23.6 - 38.3) | 0.95 (0.79-1.12) |

We examined potential sources of uncertainty in our estimates. We found higher background levels of antibody reactivity to the SARS-CoV-2 spike protein RBD in pre-pandemic samples from Juba compared to pre-pandemic samples from Boston [25] (Supplementary Figure 3). Since serological measurements from PCR-confirmed cases in Juba were not available, we could not examine whether there were also differences in post-infection antibody dynamics between the populations. However, we were able to assess the impact that different assumptions about test sensitivity had on the results. If we assumed that 60% of infections in the population were mild, we estimated 34.8% (95% CrI 30.1 - 40.2) seroprevalence (Figure 2a) and that for each reported case 94 (95% CrI 81 - 109) were unreported (Figure 2b). In contrast, if we assumed that 100% of infections were mild, we estimated 45.6% (95% CrI 35.7 - 60.6) seroprevalence (Figure 2a) and that for each reported case 123 (95% CrI 96 - 164) were unreported (Figure 2b). Regardless of assumptions, these results indicated that 98-99% of infections were unreported by August 2020.

**Figure 2.**
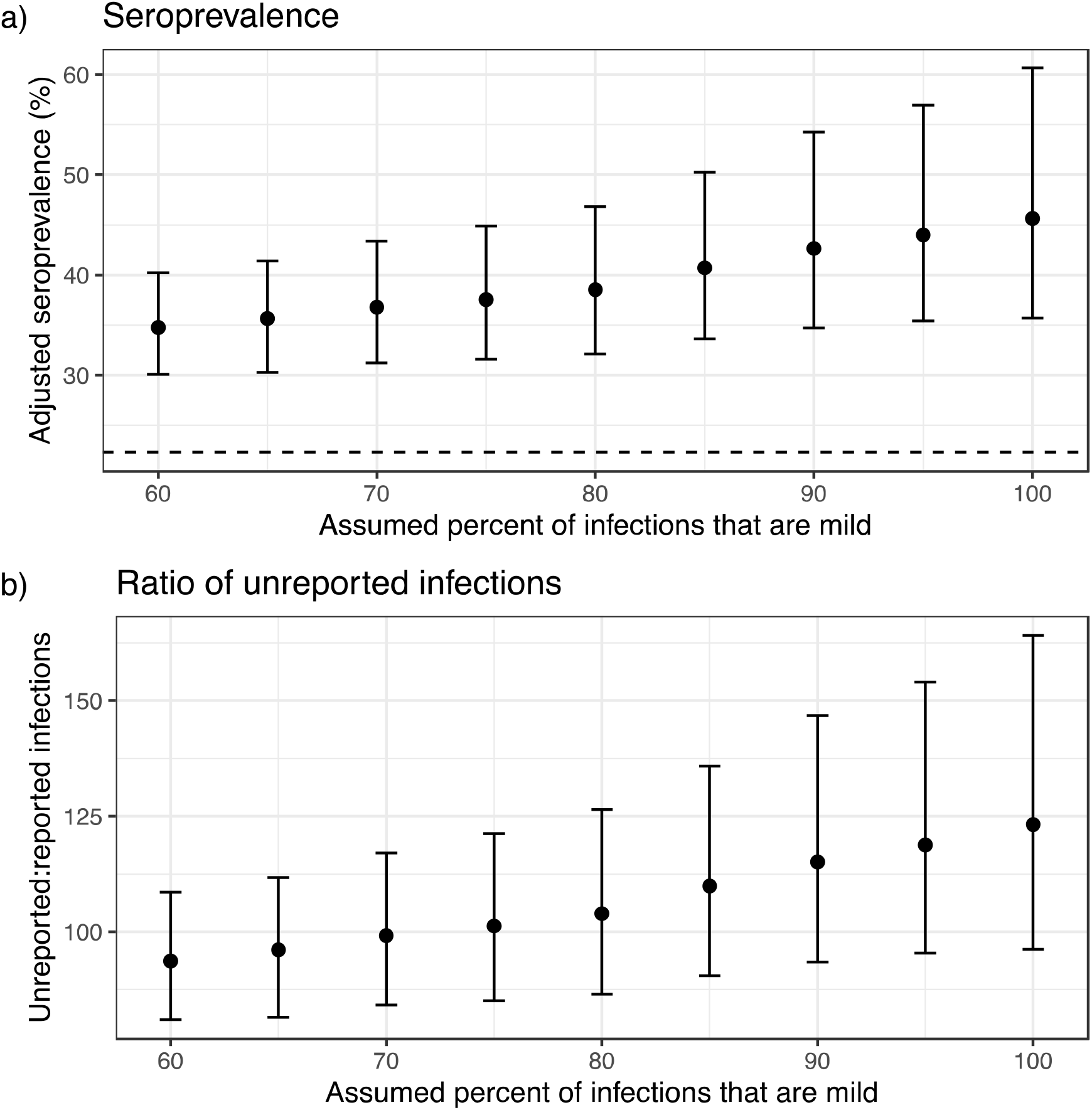
Impact of assumptions on seroprevalence and ratio of reported to unreported infections. a) Impact of assumed percent of infections that are mild on adjusted seroprevalence. Dashed line represents unadjusted seropositivity at 22.3%. b) Impact of assumed percent of infections on the ratio of implied unreported infections to reported infections, based on a total of 1,873 confirmed COVID-19 cases in Juba by August 31, 2020 and an approximate population in Juba of 510,000. Points represent mean adjusted seroprevalence or ratio of unreported infections and error bars represent 95% Credible Interval.

## Discussion

In this study we estimated that more than 1 in 3 people in Juba, South Sudan had been infected with SARS-CoV-2 by August 2020. This corresponds to 196,000 implied infections, more than 100 times the number of PCR-confirmed SARS-COV-2 infections over the same time frame. As in other sub-Saharan African populations, these results reveal that while the apparent health impacts of the COVID-19 pandemic have been lower than other parts of the world, the virus has spread extensively.

Adjusting for imperfect immunoassay performance is critical when estimating infection attack rates from serosurveys. As post-infection antibody kinetics vary by infection severity, age and prior exposures, so can test performance. Through testing pre-pandemic samples from Juba, we found that background anti-SARS-CoV-2 antibody reactivity was higher in Juba than in Boston, consistent with findings from studies conducted in other sites within sub-Saharan Africa [12,15,35,36]. We used these negative controls to estimate test specificity; however, we lack data on the post SARS-CoV-2 infection antibody kinetics and the proportion of infections that are mild or asymptomatic in the Juba population. This led to wide variation in plausible estimates of seroprevalence, as shown in sensitivity analyses. Moreover, this illustrates that immunoassay sensitivity and specificity estimates are not static and that they should be estimated for the local populations where serosurveys are being conducted.

These findings have several implications for SARS-CoV-2 control in South Sudan. At least a third of the population in Juba has been exposed to the virus, and this number has undoubtedly increased since the survey was completed in September 2020. These estimates will help the Ministry of Health and others in South Sudan weigh the costs and benefits of devoting limited resources to COVID-19 mitigation at the cost of other crucial health programs. An important open question is the extent to which SARS-CoV-2 spread and/or mitigation measures have exacerbated underlying vulnerabilities, including food insecurity, livelihoods, and co-infections such as the current measles outbreak in South Sudan [37]. Follow-up studies would be required to understand the larger impact of the epidemic in Juba, as well as in the rest of South Sudan, and better inform public health policy.

These results also have implications for SARS-CoV-2 serosurveillance more broadly. The majority of serosurveys conducted to date use sensitivity and specificity estimates directly from assay manufacturers, if they adjust seroprevalence estimates for test performance at all [5]. In many settings it may not be feasible to collect control data from local populations, but validation of different immunoassays in populations in the same region of the world where the assays are being used is critical for appropriate interpretation of study results. Moreover, our findings support previous studies that have called for the inclusion of mild and asymptomatic SARS-CoV-2 infections in assay validation datasets [38]. We and others have shown that antibody titers tend to be lower among mild and asymptomatic infections [39–44]. Thus, validation datasets comprised predominantly of severe, hospitalized cases may lead to overestimation of assay sensitivity and gross underestimation of SARS-CoV-2 seroprevalence [38].

Overall, the estimates reported in this study are comparable to SARS-CoV-2 seroprevalence estimates in Nigeria, where seroprevalence ranged from 25 to 45% depending on the population sampled [6,9,11]. Similarly, seroprevalence was 40% in public sector patients in Cape Town, South Africa [16], 12.3% (8.2 - 16.5) among asymptomatic healthcare workers in Blantyre, Malawi [13], and 25.1% among gold mine workers in Côte d’Ivoire [20]. In Addis Ababa, Ethiopia, seroprevalence among those reporting no close contact with SARS-CoV-2 infected individuals was 8.8% (5.5 - 11.6) in April [8]. Seroprevalence was lower at 4.3% (2.9 - 5.8) in blood donors in Kenya in June [7], increasing to 9.1% (7.6 - 10.8) by September [17]. These lower estimates may be due to differences in SARS-CoV-2 epidemiology in these locations, time periods, or sub-populations. Serological tests may themselves contribute to the differences; a study in Kinshasa, Democratic Republic of the Congo showed that seropositivity in health facility staff ranged from 8% to 36% depending on the serological test used [15]. Nevertheless, together these studies indicate that SARS-CoV-2 has spread widely in sub-Saharan Africa [2,3]. This conclusion is supported by a post-mortem study in Lusaka, Zambia which found that among 372 deceased individuals, 19.2% were PCR-positive for SARS-CoV-2 [45].

We also found that risk of seropositivity was lowest among participants aged 20 to 49 in Juba, in contrast to numerous other studies finding increased risk among working-aged populations [46]. This finding could accurately represent SARS-CoV-2 epidemiology in Juba if the majority of transmission occurred within households, contributing to reduced risk of spread among working individuals. Crowded living conditions among Juba’s urban population, with 31.3% of households living in shelters of 1-2 rooms and 19.5% with 4 or more members sleeping in the same room, support this hypothesis [24]. Alternatively, this finding may reflect selection bias. Since recruitment only took place during the day, working adults may have been less likely to participate, and we found that risk of seropositivity was lower among non-working participants.

This study has several limitations. As described above, our positive control data came from a cohort in Boston, USA. Thus, despite our efforts to correct for differences between the populations, we do not know how accurate our sensitivity estimates are for Juba, or anywhere on the African continent. In addition, we used a single ELISA that measured IgG antibodies targeting the RBD of SARS-CoV-2’s spike protein. Previous studies have shown variation in sensitivity and specificity of antibody assays that target different antigens [15,47], suggesting that using multiple antigens may provide a better picture of seroprevalence than a single antigen alone, particularly when validation data are not available from the local population. While the study had a standard definition for households, the study team faced challenges in implementing this strict definition, so we were unable to confidently estimate the degree to which SARS-CoV-2 infections clustered within households or to adjust for this in the regression model. In addition, many shelters originally selected using satellite imaging were empty and we had to select alternative households, which increased the time required to complete the survey. Finally, while this study was representative of the residential neighborhoods of Juba, the sample did not include more than 30,000 estimated internally displaced persons living in two of Juba’s IDP camps [48]. Nevertheless, 14.3% of households participating in the study reported to be IDPs, either living among the host-community or in another IDP site.

Despite the limitations, this is one of few population-based seroprevalence studies representative of the general population that have been conducted in sub-Saharan Africa. Furthermore, we used specificity estimates based on background antibody levels specific to the local population, adjusted seroprevalence estimates by test performance, and propagated uncertainty around test performance into our final estimates. Since the ELISA we used was quantitative, we additionally reported antibody distributions rather than seropositivity cutoffs alone (Supplementary Figure 1) and as a result it will be possible to adjust our estimates further if more accurate sensitivity data become available for this population.

In conclusion, here we present evidence that SARS-CoV-2 seroprevalence is much higher in Juba than suggested by confirmed case data alone, which is consistent with other recent serosurveys in sub-Saharan Africa. Future serosurveys in South Sudan will be helpful to confirm these findings and to examine the impact that SARS-CoV-2 spread has had on underlying vulnerabilities. Such seroprevalence studies are needed to understand the impact of the pandemic more broadly in Africa, as well as the ways to most effectively mitigate its effects. Importantly, for these efforts to be most impactful, they must be accompanied by efforts to validate serological tests in local populations.

## Data Availability

All data referred to are available within the manuscript or online in links provided.

https://github.com/HopkinsIDD/juba-sars-cov-2-serosurvey

