## Supplementary Materials for "Seroprevalence of anti-SARS-CoV-2 IgG antibodies in Juba, South Sudan: a population-based study"

#### Statistical model

Our aim was to estimate the true underlying seroprevalence of SARS-CoV-2 in the population ≥ 1 year of age in Juba, South Sudan. To that end, we estimated the probability that each participant in the serosurvey was seropositive using a Bayesian logistic regression model following Stringhini and colleagues [1] that accounts for serological test sensitivity and specificity, as well as age and sex of each participant:

$$x^i \sim Bernoulli(p_i\theta^+ + (1 - p_i) * (1 - \theta^-))$$
$$logit(p_i) = X_i\beta$$
$$x^+ \sim Binomial(n^+, \theta^+)$$
$$x^- \sim Binomial(n^-, 1 - \theta^-)$$

Here $x^i$ was the result of the IgG ELISA for each individual ($i$ = 1,…,N=1840) in the serosurvey. The probability of observing a seropositive result was a function of sensitivity, $\theta^+$ (true positive rate), and specificity, $\theta^-$ (true negative rate), in the context of the true underlying probability of seropositivity for each individual, $p_i$. This probability $p_i$ was a function of covariates $X_i$, which included the age and sex of each individual, and their coefficients $\beta$. Sensitivity, $\theta^+$, was determined using $n^+$ RT-PCR confirmed positive controls from the Boston cohort [2] (see Supplementary Figure 2 and Methods in the main text). Specificity, $\theta^-$, was determined using pre-pandemic negative controls [3], where $x^-$ tested positive. Priors on sensitivity and specificity were flat from 0 to 1 and priors on regression coefficients $\beta$ were $Normal(0,1)$.

We implemented the model in the Stan probabilistic modeling language [4] using the *rstan* package in R. We ran 5,000 total iterations, which included 4 chains with 1,500 iterations each and 250 for warm-up. The complete modeling and analysis code is available online (https://github.com/HopkinsIDD/juba-sars-cov-2-serosurvey).

### Tables

**Supplementary Table 1. Adjusted SARS-CoV-2 seroprevalence estimates for Juba.**

Primary analysis includes estimates adjusted for test performance and age and sex of the participants. The “no covariates” analysis includes estimates adjusted for test performance alone using either the subset that can be matched to age and sex data (n = 1840) or the full serological dataset (n = 2214).

| Analysis | Seroprevalence (95% CrI) |
|---|---|
| Primary (age/sex-matched data, n = 1840) | 38.5 (32.1 - 46.8) |
| No covariates (full dataset, n = 2214) | 36.2 (31.0 – 43.3) |
| No covariates (age/sex-matched data, n = 1840) | 36.9 (31.3 - 44.2) |

#### Figures

**Supplementary Figure 1. Distribution of anti-SARS-CoV-2 antibodies in the Juba population in 2015 before the pandemic (n = 104) and during the survey (n = 2214).**

Histogram of IgG titers in 2015 before the pandemic (top) and in 2020 during the survey (bottom). The purple dashed line indicates the maximum value detected in any pre-pandemic sample (0.32 ug/mL), which is used as the seropositivity cutoff.

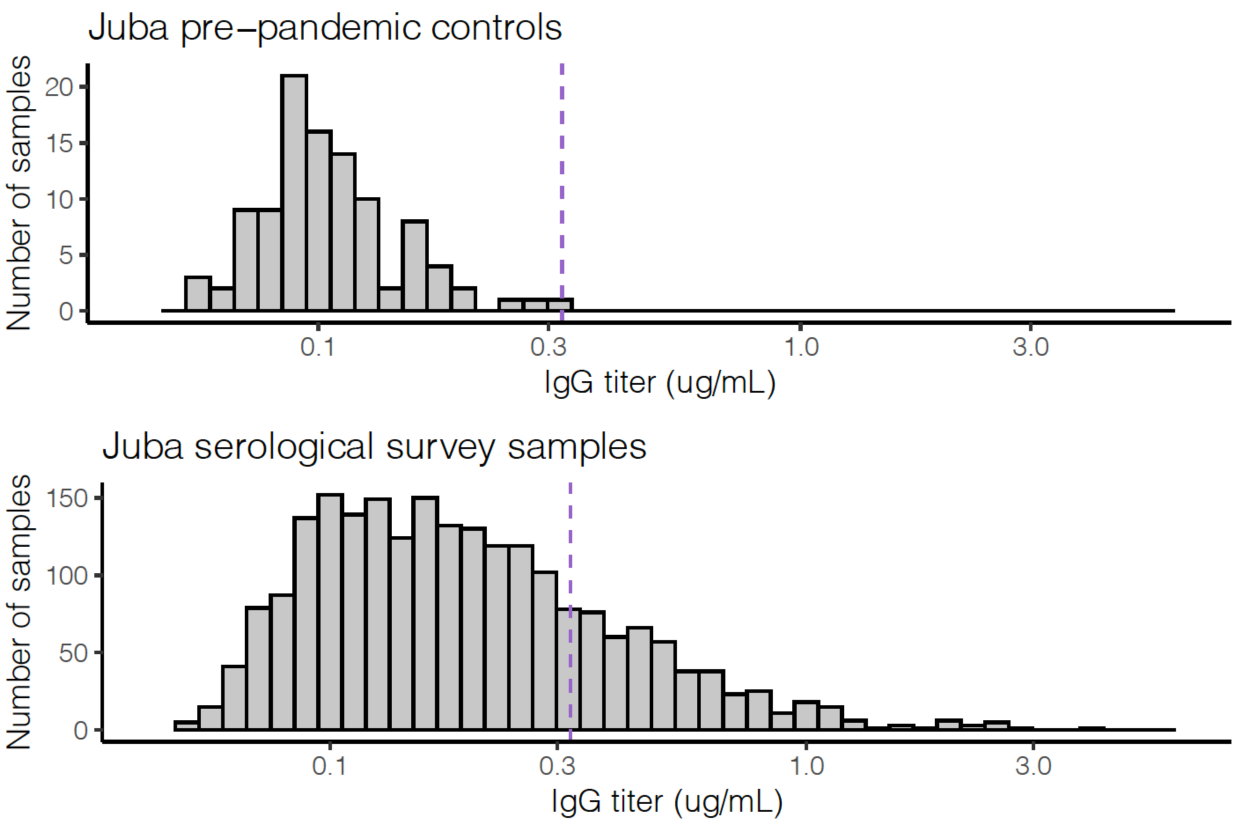

**Supplementary Figure 2. Antibody dynamics in the Boston cohort.**

Antibody titers at various time points post symptom onset stratified by severity and compared to pre-pandemic controls. Black points and smoothed trajectories for days 0 to 70 represent data from Iyer, Jones *et al*. 2020. Purple points and trajectories represent additional data from mild PCR-confirmed COVID-19 cases (as described in the Methods section of the manuscript). Data from individuals who died are not included.

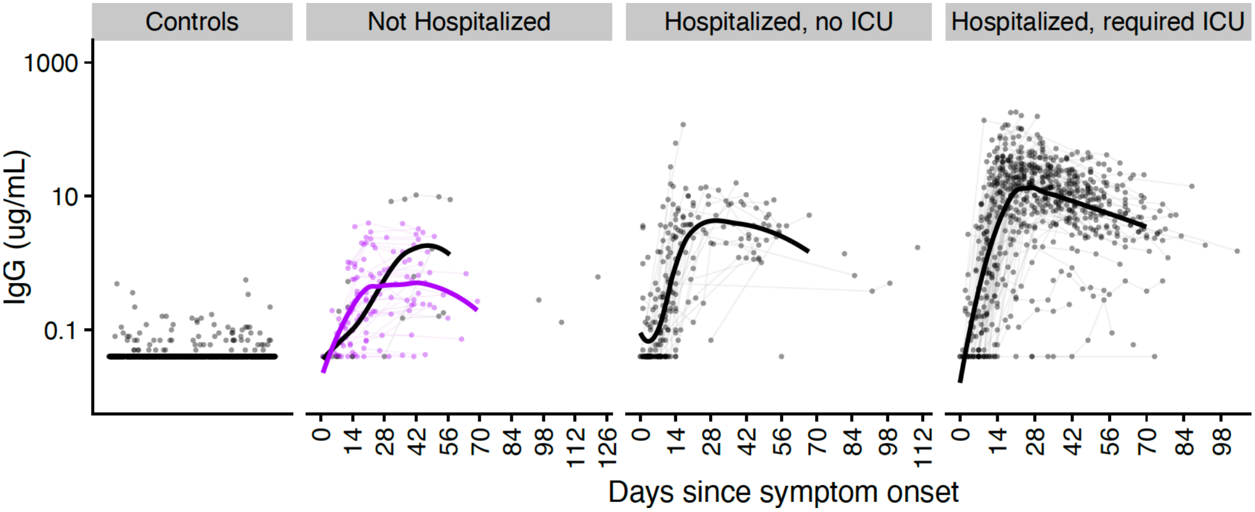

**Supplementary Figure 3. Antibody distributions in pre-pandemic negative controls from populations in Boston, USA (n = 1548) and Juba, South Sudan (n = 104).**

Each point represents an individual. The dashed line represents the limit of detection of the serological test. Boston data collected before the pandemic are shown in green and represent a combination of healthy adults seen at the Massachusetts General Hospital travel clinic, patients undergoing routine serology testing at Massachusetts General Hospital, and patients presenting with a known febrile illness. The green line at the limit of detection indicates that the vast majority of samples had such low background reactivity that they fell below this limit. Juba data collected in 2015 are shown in purple; none of the Juba samples fell below the limit of detection.

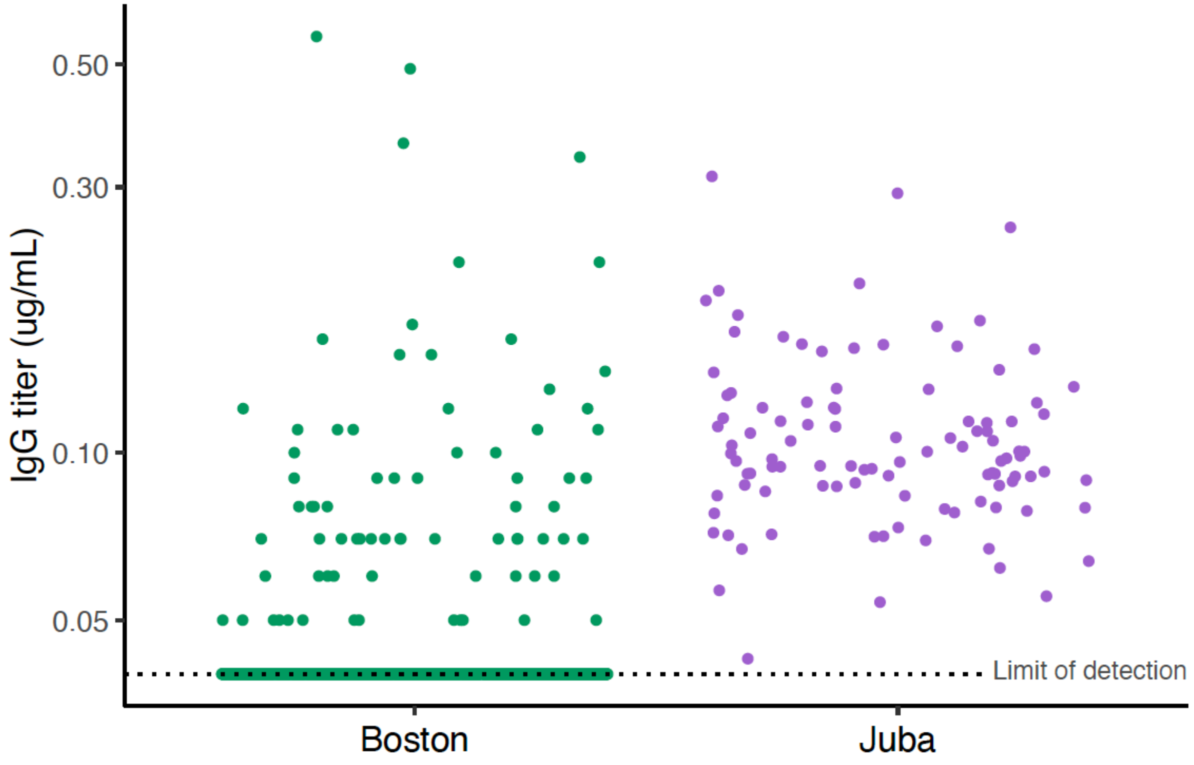
